# Effects Of Mindfulness On Sleep Quality, Anxiety, Depression and Stress In Migraine

**DOI:** 10.1101/2020.07.20.20158212

**Authors:** Shana AB Burrowes, Olga Goloubeva, Kristen Stafford, Patrick F. McArdle, Madhav Goyal, B. Lee Peterlin, Jennifer A. Haythornthwaite, David A. Seminowicz

**Affiliations:** Boston University School of Medicine, Section of Infectious Diseases, Department of Medicine, Boston MA, USA, University of Maryland Baltimore Department of Neural and Pain Sciences, School of Dentistry, Center to Advance Chronic Pain Research, Department of Epidemiology and Public Health, School of Medicine, Baltimore, MD, USA; University of Maryland Baltimore, University of Maryland Greenebaum Comprehensive Cancer Center, Baltimore, MD, USA; University of Maryland Baltimore, Department of Epidemiology and Public Health, School of Medicine Baltimore, MD, USA; University of Maryland Baltimore, Department of Medicine, School of Medicine, Baltimore, MD, USA; Johns Hopkins University School of Medicine, Department of Medicine, Division of General Internal Medicine, Baltimore, MD, USA; Penn Medicine Lancaster General Health, Neuroscience Institute, Lancaster, PA, USA; Johns Hopkins University School of Medicine, Department of Psychiatry and Behavioral Sciences, Baltimore, MD, USA; University of Maryland Baltimore, Department of Neural and Pain Sciences, School of Dentistry, Center to Advance Chronic Pain Research, Baltimore, MD, USA

**Keywords:** migraine, sleep, psychosocial, meditation, nonpharmacological

## Abstract

**Objective:** To examine the change over time in sleep quality and psychosocial outcomes from the MRI Outcomes for Mindfulness Meditation Clinical Trial, assess how these mediated treatment response (50% reduction in headache frequency post-intervention) and examine the relationship between baseline values and treatment response.

**Methods:** This is a secondary analysis of a randomized placebo-controlled trial with single masking. Patients were recruited from the community, headache clinics and primary care offices in Baltimore Maryland. Eligible patients were aged 18-65 and met the International Classification of Headache Disorders criteria for episodic migraine (4-14 headache days in 28 days).The trial (primary outcomes previously reported) included 98 episodic migraine patients randomized to either enhanced (MBSR+) or stress management for headache (SMH). Follow-up visits occurred at 10, 20 and 52 weeks post baseline. Intervention groups met separately for two hours weekly for 8 weeks and then biweekly for an additional 8 weeks. MBSR+ had an additional half-day retreat held between weeks 6 and 8.

**Results:** There was a significant improvement in sleep quality from baseline to post-intervention (p=0.0025), in both groups. There were no significant change from baseline or between groups in anxiety, depression and stress. There was also no significant association between baseline scores and treatment response. Mediation analysis showed a significant indirect effect for sleep, anxiety, stress, and depression on treatment response, ranging from 6-8%.

**Conclusions:** The findings indicate that in episodic migraine, treatment response to MBSR+ compared to SMH was mediated by small, but significant improvements in sleep, anxiety, stress, and depression.

## Introduction

Sleep disturbances^1,2^ and psychosocial conditions such as anxiety and depression are more prevalent in migraine sufferers than the general population^3^, and these co-morbid conditions complicate treating migraine. Current migraine therapies either reduce pain or prevent migraines.^4^ However, relief from migraine is a combination of the physical components (e.g. pain, nausea), and the affective factors, both of which are targeted by non-pharmacological interventions such as mindfulness based stress reduction (MBSR).^5,6^

MBSR and other mindfulness therapies have been associated with improvement in pain and function^5,7,8^ and reduction in headache frequency in headache patients.^6,9^ Reductions in stress, anxiety and depression have been extensively reported in healthy and patient populations.^10-12^ Although no studies that show the mediating effect of psychosocial factors in migraine, evidence in other populations indicate that individual psychological factors play a role in treatment success.^13-16^

MBSR is likely useful in reducing headache pain and psychosocial co-morbidities in migraineurs. Patients may also have better sleep quality, related to MBSR and or improvement in headaches. This secondary data analysis from our clinical trial^17^ assessed enhanced MRSR (MBSR+) as a comprehensive therapy for migraine. We hypothesized that a) compared to SMH, patients randomized to MBSR+ have greater improvement in sleep quality, anxiety, depression and stress symptoms over time compared to an active control treatment arm, b) baseline sleep quality, anxiety, depression and stress symptoms predict treatment response to MBSR+ and c) sleep quality, anxiety, depression and stress symptoms are mediators of the effect of MBSR+ on treatment response.

## Methods

### Clinical Trial Design

This secondary analysis utilized data from the MRI Outcomes of Mindfulness Meditation for Migraine clinical trial (NCT02133209).^17^ Participants were enrolled from June 2014 to February 2017. The primary objective of the clinical trial was to determine the short and long term efficacy of MBSR+ on headache frequency, brain structure and function. This was a randomized placebo-controlled trial with single masking. All migraine patients were episodic migraineurs (EM) who were randomly assigned to receive either MBSR+ or SMH. Patients were randomized 1:1 to either treatment in pre-specified blocks, using a web-based randomization system. Randomization was stratified by the presence or absence of another chronic pain disorder and by headache frequency: 4-8 (low frequency) or 9-14 (high frequency) headache days per 28 days. The clinical trial also enrolled 30 healthy pain free controls, matched to the migraine patients on age (±5 years), sex, body-mass index (BMI) (±5), education (college and no college) and race. Data from healthy controls were not utilized in these analyses. Each treatment was administered weekly for 8 weeks with an additional half day retreat for the MBSR+ arm and then both treatments were administered for an additional 4 sessions over 8 weeks. Full inclusion and exclusion criteria have been previously reported.^17^

### Outcome Measures

Patients were assessed at three time points: baseline (pre-randomization), mid-intervention (week 10) and post-intervention (week 20). The mid-intervention assessment occurred following the first 8 weeks of intervention and the post-intervention assessment occurred following the second 8 weeks.

#### Headache Frequency: Assessment of treatment responders

Daily headache diaries were completed electronically (online via a link sent through email) and collected detailed information on headaches over a 28-day period (e.g. pain intensity, headache duration and other symptoms). Headache diaries were used to ascertain clinical endpoints including headache frequency at baseline and over time. Given the nature of headache diaries not all patients completed a full 28 days. To address this, we calculated a proportion for everyone, by dividing the number of headache days reported by the total number of diary days collected in that period. For any given individual the maximum denominator was 28. Additionally, we multiplied the proportion by 28 to get a continuous variable for headache days to utilize in final models. To calculate the responders in the sample we used the above proportion and the standardized headache days variable and calculated the difference between baseline and the post-intervention time point. Treatment responders were defined as achieving at least a 50% reduction in headache frequency from baseline to post-intervention (week 20).

#### Sleep quality and psychosocial questionnaires

Sleep quality was assessed using the Pittsburgh Sleep Quality Index (PSQI) questionnaire^18^, a 23 item scale that assesses the quality of sleep over the last month with possible scores ranging from 0-21.^18^ Anxiety was assessed using the Generalized Anxiety Disorder (GAD-7) questionnaire, ^19,20^ a 7 item questionnaire that measures common symptoms of anxiety with scores ranging from 0-21. Depression was measured with the Patient Health Questionnaire-9 (PHQ-9),^21^ a nine item questionnaire for depressive symptom severity over the last two weeks with scores ranging from 0-27. Stress was assessed with the Perceived Stress Scale (PSS), ^22,23^ a 10 item questionnaire that measures an individual’s perceived stress to everyday life situations over the last month with scores ranging from 0-40. Questionnaire scores were also categorized according to validated cut points. PSQI total scores ≥5 indicate poor sleep quality. GAD-7 was categorized into four groups: 0-4 (no anxiety symptoms), 5-9 (mild anxiety symptoms), 10-14 (moderate anxiety symptoms), 15≤ (severe anxiety symptoms). Similarly, PHQ-9 was separated into 5 categories: (0-4, 5-9, 10-14, 15-19 and ≥20) to represent no depression symptoms, mild, moderate, moderately severe and severe depression symptoms. PSS was categorized into three groups where 0-13 is average, 14-26 is moderate stress and high stress is 27-40. Finally, patients were assessed as either having symptoms or not for all of the above questionnaires (e.g. depression symptoms yes/no).

### Statistical Analysis

#### Demographic and Clinical Variables

Distributions of baseline patients’ characteristics between those randomized to MBSR+ and SMH and differences between treatment responders and non-responders were assessed utilizing Chi-square, t-tests, and nonparametric Wilcoxon and Kruskal-Wallis tests. Baseline characteristics included headache frequency, presence of an additional idiopathic pain condition, education, age, sex, race and BMI. In addition to sleep quality and psychosocial factors, the following clinical headache characteristics were examined: headache days, mean headache pain and headache impact test (HIT-6) scores. All analyses utilized intention to treat (ITT) analysis, therefore patients were analyzed as randomized.

#### Change in sleep quality and psychosocial symptom scores over time

Exploratory data analysis examined the distributions of baseline scores and the change in scores over time (baseline, 10 and 20 weeks) of all questionnaires. Wilcoxon, ANOVA and repeated measures ANOVA were used to assess differences in group, time and group by time interactions. Several regression diagnostic measures were utilized to examine the linearity of variables as outcomes and to assess the correlation of these variables as predictors. Using variance inflation factor (VIF), Cook’s D, plots of both the raw and scaled residuals and predicted values we examined the assumptions of normality, linearity and independence of these variables as both outcome and predictive measures. Given the skewed nature of PSQI, GAD-7 and PHQ-9 we also assessed the utility of log transformation on these factors. Using mixed effects model diagnostics it was determined to use the untransformed scores in all final models.

A random intercept was included in the final model for depression only as this was the only outcome where this approach improved the model fit. All models were corrected for multiple comparisons, utilized an unstructured covariance and assessed differences between MBSR+ and SMH groups, change from baseline and group by time interactions. Models were assessed using the Akaike Information Criterion (AIC) as well as the impact of additional variables on the standard error.

#### Baseline sleep quality and psychosocial scores and the relationship with treatment response

PSQI, GAD-7, PHQ-9 and PSS scores are known to be related and initial analyses assessed the statistical correlation using Spearman’s Rho as well as their impact on each other by examining collinearity diagnostics. Rho values equal to or more then 0.5 as well as a p value ≤0.05 were assessed further. We assessed condition index, variance inflation factor, tolerance and proportion of variance values. It was determined that each predictor would be modeled separately to assess their individual association with treatment response.

Individual models assessed baseline sleep and psychosocial variables as the predictor, adjusting for treatment group. Confounders of interest at baseline were BMI, education, race, sex, age, idiopathic pain condition and HIT-6. We also assessed the role of baseline mindfulness and pain catastrophizing as potential confounders. Final models were assessed as described above. All statistical analyses were conducted using SAS (v.9.4, SAS Institute Inc. Cary, NC). Testing was two-sided and done at the 0.05 level of significance.

#### Mediation of Treatment Response by Change in Psychosocial Scores over Time

We examined the mediation of treatment effects (MBSR+ vs. SMH) on treatment response through sleep, anxiety, depression and stress. To assess the presence of mediation via these factors we employed mediation analysis methods for binary outcomes and multiple mediators as outlined by VanderWeele^24^ and VanderWeele and Vansteelandt.^25^ Figure 1 depicts the directed acyclic graph (DAG) that guided the development of the final models. The weighting method developed by VanderWeele and Vansteelandt utilizes an inverse probability of treatment weighting approach to estimate the direct and indirect effects even when mediators impact each other. ^25^

**Figure 1:**
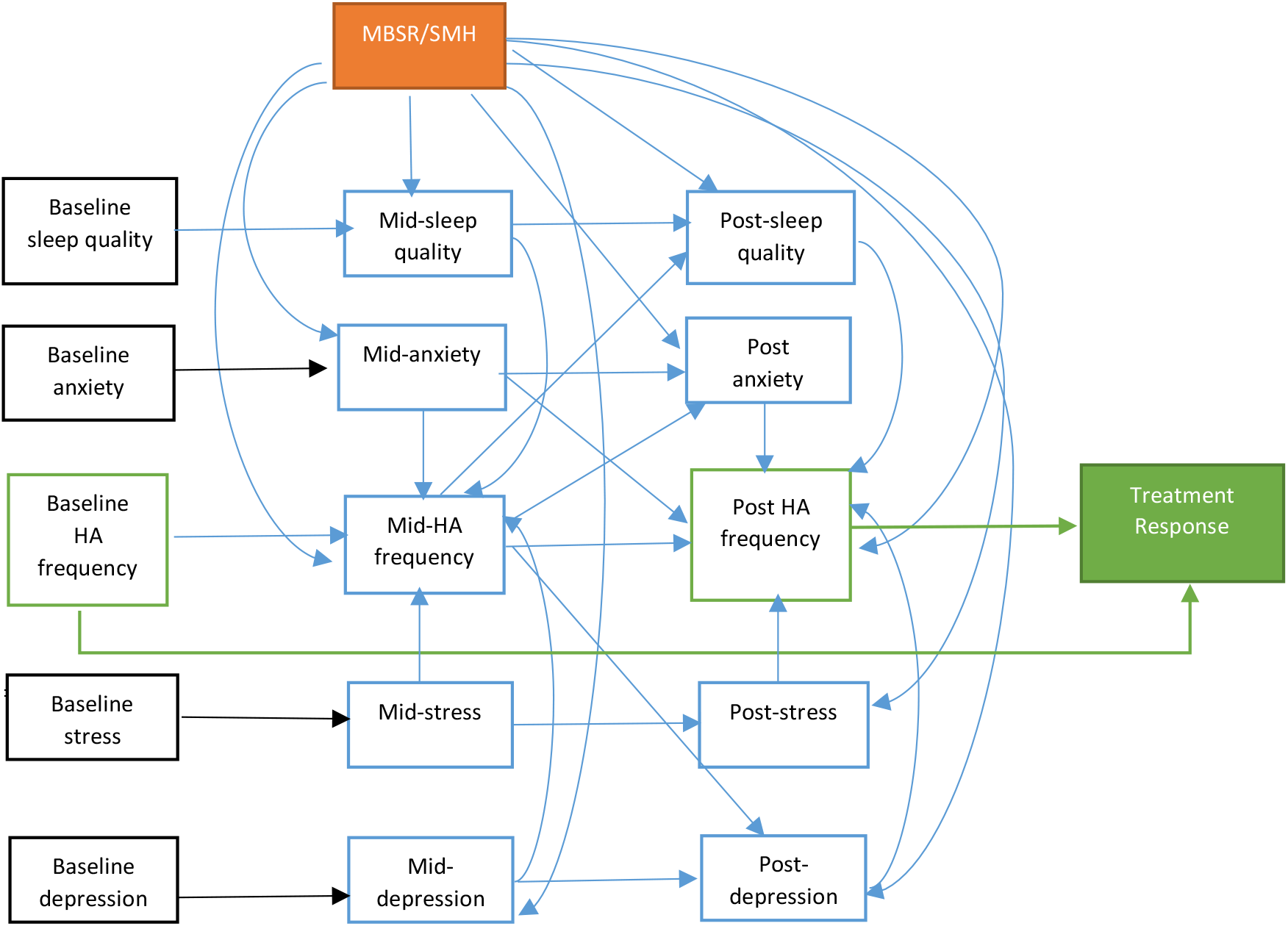
Causal diagram to represent plausible mediation relationship between MBSR+ and treatment response. Arrows assume direction of causal relationships. Model assumes no unmeasured confounding. Black boxes indicate baseline covariates. Solid black arrows represent adjusted confounding. The orange box indicates the randomized treatment MBSR+ vs. SMH. The solid green box is the outcome which is treatment response. This variable is created from baseline and post-intervention headache frequency (green outlined boxes) Blue boxes represent possible mediators of the relationship between exposure and outcome. Baseline covariates were measured prior to randomization and therefore are not affected by the treatment. Mid-point (10 weeks post baseline) sleep quality, anxiety, depression and stress were measured at the scan visit directly after the completion of the first block of treatment. Headache diaries were completed for 28 days post the first block of treatment and therefore are captured after the psychosocial measurements. Post-treatment (20 weeks post baseline) psychosocial measurements were taken at the scan visit directly after the final treatment block had been completed. These visits usually occur over a two-week period post study completion. Headache diaries are completed for 28 days post treatment.

This approach estimates three weighted averages which correspond to the following counterfactuals: E[Treatment response_a\****M***a*_], E[Treatment response_a***M***a_] and E[Treatment response_a***M***a*_] where a= MBSR+ and a*= SMH and M = mediator vector. Each is calculated in a three step process. For E[Treatment response_a\****M***a*_], the first step fits a logistic regression model using a mediator vector (this included psychosocial factors of interest and headache frequency reported at 10 weeks) and baseline covariates to estimate predicted probabilities of P(SMH|c_i_). Secondly a weighted average of the predicted values for subjects is calculated by P(A=SMH)/P(A=SMH|c_i_). The final step utilizes the weight in a regression model to estimate the value of E[Treatment response_a\****M***a*_]. The process is repeated for each counterfactual component. The direct effect is given by *E*[*Treatment response*_*a****M****a\**_]−*E*[*Treatment response*_*a\*****M****a\**_] and the indirect effect: *E*[*Treatment response*_*a****M****a*_] − *E*[*Treatment response*_*a****M****a\**_].

All models assessed each mediator separately, adjusting for baseline values of the remaining psychosocial variables. Exposure-mediator (treatment group*psychosocial factor) and mediator-mediator interactions were assessed in all models, however these models did not differ significantly from those without interaction. Given that no interactions, the natural direct effect is assumed equivalent to the controlled direct effect. To calculate standard errors and 95% confidence intervals for each estimate a bootstrapping approach was employed. This process generated 1000 samples with replacement after which counterfactual estimates and confidence intervals were generated for all samples. Confidence intervals that did not include the null were significant. Analyses were conducted using SAS (v.9.4, SAS Institute Inc. Cary, NC).

### Standard Protocol Approvals, Registrations and Patient Consents

The study was approved by the Johns Hopkins School of Medicine and University of Maryland Baltimore Institutional Review Boards. All patients provided written informed consent. Secondary analysis of data from registered clinical trial NCT02133209.

### Data availability statement

All data is available upon request.

## Results

### Description of baseline demographic and clinical measures in episodic migraine patients

Of the 583 patients screened, 122 completed baseline assessments and 98 were eligible and randomized to receive either MBSR+ (n = 50) or SMH (n = 48). All 98 patients were included in the analyses examining change in sleep and psychosocial metrics over time. Treatment response analyses were reduced to the 95 patients with baseline and post-intervention follow-up data that was needed to assess the outcome.

Migraine patients randomized to MBSR+ or SMH were of comparable age, race, sex, BMI and educational attainment. Both groups were predominantly female, White, college educated with a median age of 36 (Table 1). Clinical characteristics, particularly those related to migraine, were also similar between the randomized groups. The proportion of headache days ranged from 0.11-0.5 in both groups with an average value of 0.25 in the SMH group and 0.29 in the MBSR+ group. Overall 29% of patients reported poor sleep quality, 13% of all patients met criteria for anxiety symptoms 20% for depression symptoms and 39% for stress. There was no significant difference in the proportions between MBSR+ and SMH groups (data not shown).

**Table 1:** **Baseline clinical and demographic variables in episodic migraine patients randomized to either MBSR+ or SMH**

| Variable | MBSR+(n=50) | SMH (n=48) | p value<sup>a</sup> |
| --- | --- | --- | --- |
| Sex/ n (%) |  |  |  |
| Male | 3 (33.33) | 6 (66.67) | 0.31 |
| Female | 47 (52.81) | 42 (47.19) |  |
| Age/median (range) | 36 (18-65) | 35.5 (21-63) | 0.44 |
| Race/n (%) |  |  |  |
| Black | 10 (58.82) | 7 (41.18) | 0.79 |
| White | 35 (49.3) | 36 (50.7) |  |
| Other | 4 (44.44) | 5 (55.56) |  |
| Unknown | 1(100) | 0 |  |
| Education/ n (%) |  |  |  |
| Some College or Less | 13 (65) | 7 (35) | 0.16 |
| College and Above | 37 (47.44) | 41 (52.56) |  |
| BMI/median (range) | 27.30 (15.66-47.44) | 25.71 (16.63-45.45) | 0.79 |
| Migraine Qualities at Baseline |  |  |  |
| Low Frequency (4-8 days)/n (%) | 25 (50) | 25 (50) | 0.84 |
| High Frequency (9-14 days)/n (%) | 25 (52.08) | 23 (47.92) |  |
| Headache Days/median<br>(range) | 8 (3-14) | 7 (3-14) | 0.83 |
| Headache Pain/mean±SD | 4.72±1.65 | 4.31±1.57 | 0.22 |
| **HIT-6 Total mean±SD** | 61±5.83 | 60.85±5.586 | 0.52 |
| **Idiopathic Pain/n (%)** |  |  |  |
| Yes | 15 (53.57) | 13 (46.43) | 0.75 |
| No | 35 (50) | 35 (50) |  |
| **Preventative Migraine Medication Use<sup>b</sup>/n (%)** |  |  |  |
| Yes | 11 (73.33) | 4 (26.67) | 0.06 |
| No | 39 (46.99) | 44 (53.01) |  |
| **Sleep quality and psychosocial measures/**<br>**Median (range)** |  |  |  |
| PSQI | 3.5(0- 11.67) | 4.67 (0-14) | 0.77 |
| GAD-7 | 1 (0-16) | 1.5 (0-16) | 0.08 |
| PHQ-9 | 2 (0-13) | 3 (0-7) | 0.07 |
| PSS | 9 (2-32) | 13 (1-26) | 0.18 |

Abbreviations: HIT-6, headache impact test; PSS, perceived stress score; GAD-7, generalized anxiety disorder; PHQ-9, patient healthy questionnaire; PSQI, Pittsburgh sleep quality index.
<sup>a</sup>p values calculated from T tests, Chi Square, Wilcoxon, Kruskal Wallis tests.
<sup>b</sup>Preventative medications cover four categories (list categories here) of medications which have been combined in this table.

### Change over time in MBSR+ and SMH groups

#### Sleep Quality

Sleep quality results show that though patients randomized to MBSR+ on average had lower scores than those randomized to SMH, the difference between the groups was not significant (Table 2). However, the change in sleep quality across groups over time was significant. From baseline to week 10 there was a decline of −0.13 (95% CI: −0.70, 0.45) and further decline from baseline to 20 weeks −0.67 (95% CI: -1.10, −0.24) (Table 2).

**Table 2:** **Adjusted analyses assessing effects of group and time on psychosocial factors in episodic migraine patients**

| Variable | Mean Difference (95% Confidence Interval) | p value<sup>a</sup> |
| --- | --- | --- |
| **Sleep Quality (PSQI)<sup>b</sup>** |  |  |
| **Group effect** |  |  |
| MBSR+ vs. SMH | -0.42 (-1.39, 0.55) | 0.39 |
| **Time effect** |  |  |
| Δ 10 weeks | -0.13 (-0.70, 0.45) | 0.66 |
| Δ 20 weeks | -0.67 (-1.10, -0.24) | **0.0025** |
| **Anxiety (GAD-7)<sup>c</sup>** |  |  |
| **Group Effect** |  |  |
| MBSR+ vs. SMH | -0.13 (-0.71, 0.46) | 0.67 |
| **Time Effect** |  |  |
| Δ 10 weeks | -0.20 (-0.66, 0.25) | 0.38 |
| Δ 20 weeks | 0.14 (-0.26, 0.55) | 0.48 |
| **Depression (PHQ-9)<sup>d</sup>** |  |  |
| **Random effects variance parameter** |  |  |
| Intercept | 2.62 (1.85, 3.98) |  |
| Residual | 2.23 (1.83, 2.78) |  |
| **Group Effect** |  |  |
| MBSR+ vs. SMH | -0.14 (-0.88, 0.61) | 0.72 |
| **Time Effect** |  |  |
| Δ 10 weeks | 0.35 (-0.09, 0.80) | 0.12 |
| Δ 20 weeks | -0.11 (-0.57, 0.35) | 0.64 |
| **Stress (PSS)<sup>e</sup>** |  |  |
| **Group Effect** |  |  |
| MBSR+ vs. SMH | -0.93 (-2.49, 0.62) | 0.24 |
| **Time Effect** |  |  |
| Δ 10 weeks | 0.09 (-0.84, 1.03) | 0.85 |
| Δ 20 weeks | -0.49 (-1.65, 0.67) | 0.40 |

Abbreviations: PSS, perceived stress score; GAD-7, generalized anxiety disorder; PHQ-9, patient healthy questionnaire; PSQI, Pittsburgh sleep quality index.
<sup>a</sup>p value from linear mixed model, FWE corrected
<sup>b</sup>Model adjusted for stress, BMI and headache pain
<sup>c</sup>Model adjusted for stress and depression.
<sup>d</sup>Model adjusted for anxiety, headache impact score and additional idiopathic pain condition. Model includes random intercept.
<sup>e</sup>Model adjusted for depression, anxiety, sleep and headache days

#### Psychosocial factors

Although patients randomized to MBSR+ had lower scores across all metrics, the difference between groups was not statistically significant (Table 2). With the exception of anxiety symptoms (an increase at 20 weeks), the scores for all other metrics had declined at 20 weeks. However, these were not significant (Table 2).

### Baseline sleep quality, anxiety, depression and stress symptom scores and the association with treatment response

Table 3 provides a description of the patients included in this analysis. There was no significant association between any baseline symptom score and treatment response. However, they all showed similar relationships with treatment success where higher scores were associated with increased odds of treatment response. The OR associated with baseline PSQI was 1.1, 95% CI: 0.94, 1.3). This pattern was consistent across GAD-7 (OR =1.11, 95% CI: 0.95, 1.30), PHQ-9 (OR= 1.13, 95% CI: 0.92, 1.38) and PSS (OR= 1.03, 95% CI: 0.96, 1.11) (Table 4). There was no interaction between intervention arm and baseline psychosocial score (continuous or categorical). Therefore, results are not stratified by intervention arm.

**Table 3:** **Baseline clinical and demographic variables in episodic migraine patients by treatment responder status**

| Variable | Treatment |  | p value<sup>a</sup> |
| --- | --- | --- | --- |
|  | Responder |  |  |
|  | Yes (n=37) | No (n=58) |  |
| **Sex (%)** |  |  |  |
| Male | 2 (25) | 6 (75) | 0.48 |
| Female | 35 (40.23) | 52 (59.77) |  |
| **Age/median (range)** | 35 (22-65) | 37 (18-63) | 0.77 |
| **Race/n (%)** |  |  |  |
| Black | 8 (50) | 8 (50) | 0.06 |
| White | 22 (31.88) | 47 (68.12) |  |
| Other | 6 (66.67) | 3 (33.33) |  |
| Unknown | 1 (100) | 0 |  |
| **Education/ n (%)** |  |  |  |
| Some College or Less | 11 (57.89) | 8 (42.11) | 0.06 |
| College and Above | 26 (34.21) | 50 (65.79) |  |
| **BMI/median (range)** | 28.64 (16.75-47.44) | 25.03 (15.66-45.45) | 0.05 |
| **Migraine Qualities at Baseline** |  |  |  |
| Low Frequency (4-8 days)/n (%) | 16 (33.33) | 32 (66.67) | 0.26 |
| High Frequency (9-14 days)/n (%) | 21 (44.68) | 26 (55.32) |  |
| Headache Days/median (range) | 9 (5-13) | 8 (3-14) | 0.15 |
| Headache Pain/mean±SD | 4.74±1.38 | 4.35±1.59 | 0.22 |
| **HIT-6 Total mean±SD** | 60.59± 5.86 | 61± 5.40 | 0.73 |
| **Idiopathic Pain/n (%)** |  |  |  |
| Yes | 15 (55.56) | 12 (44.44) | **0.04** |
| No | 22 (32.35) | 46 (67.65) |  |
| **Preventative Migraine Medication** |  |  |  |
| **Use<sup>b</sup>/n (%)** |  |  |  |
| Yes | 8 (57.14) | 6 (42.86) | 0.13 |
| No | 29 (35.8) | 52 (64.2) |  |
| **Sleep quality and psychosocial** |  |  |  |
| **measures/ Median (range)** |  |  |  |
| PSQI | 4.67 (1.17-11.67) | 3.5 (0-14) | 0.07 |
| GAD-7 | 1 (0-16) | 1 (0-16) | 0.63 |
| PHQ-9 | 3 (0-13) | 2 (0-10) | 0.38 |
| PSS | 12 (2-28) | 11.5 (1-32) | 0.28 |

Abbreviations: HIT-6, headache impact test; PSS, perceived stress score; GAD-7, generalized anxiety disorder; PHQ-9, patient healthy questionnaire; PSQI, Pittsburgh sleep quality index.
<sup>a</sup>p values calculated from T tests, Chi Square, Wilcoxon, Kruskal Wallis tests.
<sup>b</sup>Preventative medications cover four categories of medications which have been combined in this table.

**Table 4:** **Adjusted models to examine the association between baseline sleep quality, psychosocial scores and treatment response**

| Variable | Odds Ratio (95% CI)<sup>a</sup> | p value<sup>a</sup> |
| --- | --- | --- |
| **Sleep Quality <sup>b</sup>** |  |  |
| PSQI | 1.10 (0.94, 1.30) | 0.24 |
| **Anxiety <sup>c</sup>** |  |  |
| GAD-7 | 1.11 (0.95, 1.30) | 0.19 |
| **Depression <sup>c</sup>** |  |  |
| PHQ-9 | 1.13 (0.92, 1.38) | 0.25 |
| **Stress <sup>c</sup>** |  |  |
| PSS | 1.03 (0.96, 1.11) | 0.39 |

Abbreviations: GAD-7, generalized anxiety disorder; PHQ-9, patient healthy questionnaire; PSS, perceived stress score; SMH, Stress Management for Headache
<sup>a</sup> p value from adjusted logistic regression model
<sup>b</sup> Model adjusted for randomization group (MBSR+ vs. SMH), race and BMI
<sup>c</sup> Models adjusted for randomization group (MBSR+ vs. SMH), additional idiopathic pain condition and BMI

### Mediation of Treatment Response by Change in Psychosocial Scores over Time

The total causal effect, that is the total difference in treatment response between those randomized to MBSR+ and SMH is 28-29% (Table 5). The direct effect of MBSR+, that is, the effect of MBSR+ independent of any other pathways ranged from 19-22% (Table 6).The direct effect for the sleep model was 0.19 (95% CI 0.18, 0.2), indicating that 19% of the effect of MBSR+ is through pathways which do not include sleep. The indirect effect, the effect of MBSR+ mediated through pathways that included sleep, was 0.06 (95% CI: 0.05, 0.08). This shows that 6% of the effect of MBSR+ was through the improvement of sleep. This suggests that an additional 6% in treatment response could be seen in those randomized to SMH if their sleep quality had improved to the same extent as those who received MBSR+ (Table 6). Similar results were observed in models for anxiety, depression and stress symptoms (Table 6).

**Table 5:**
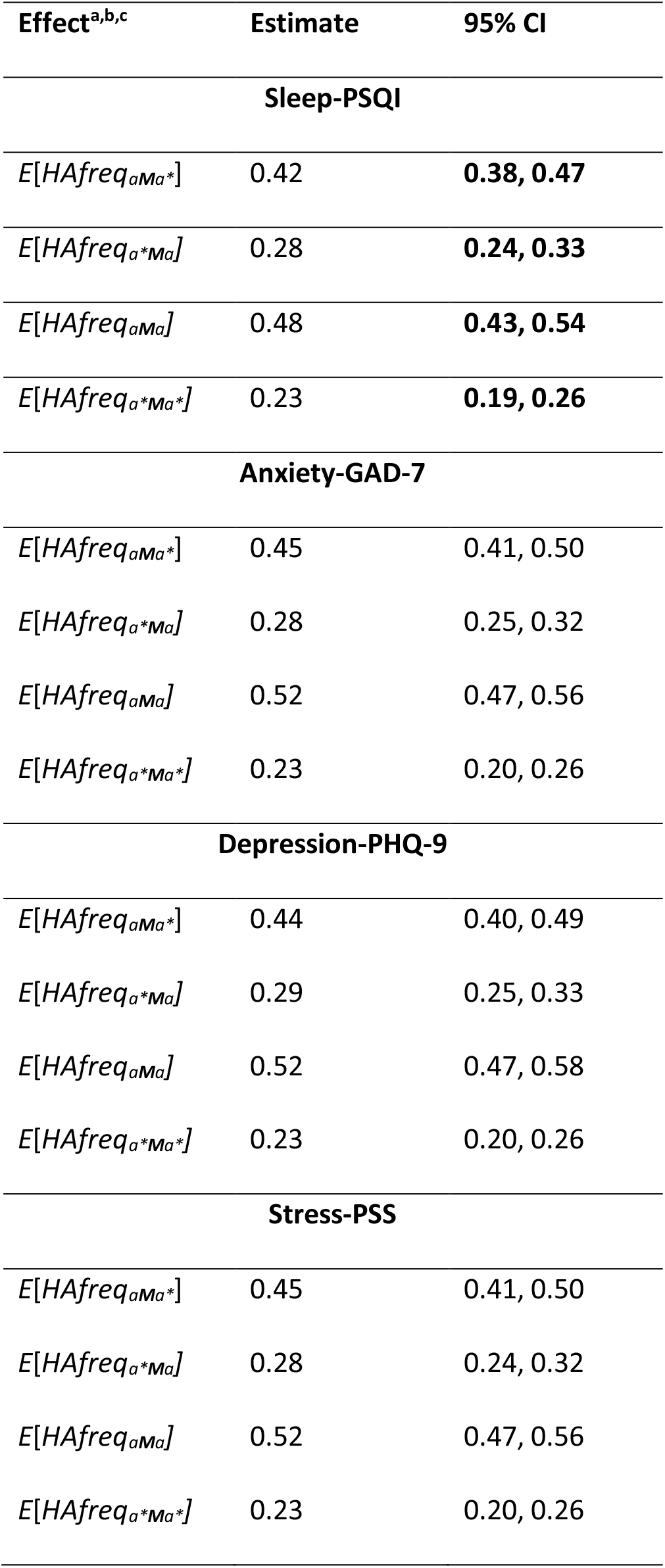
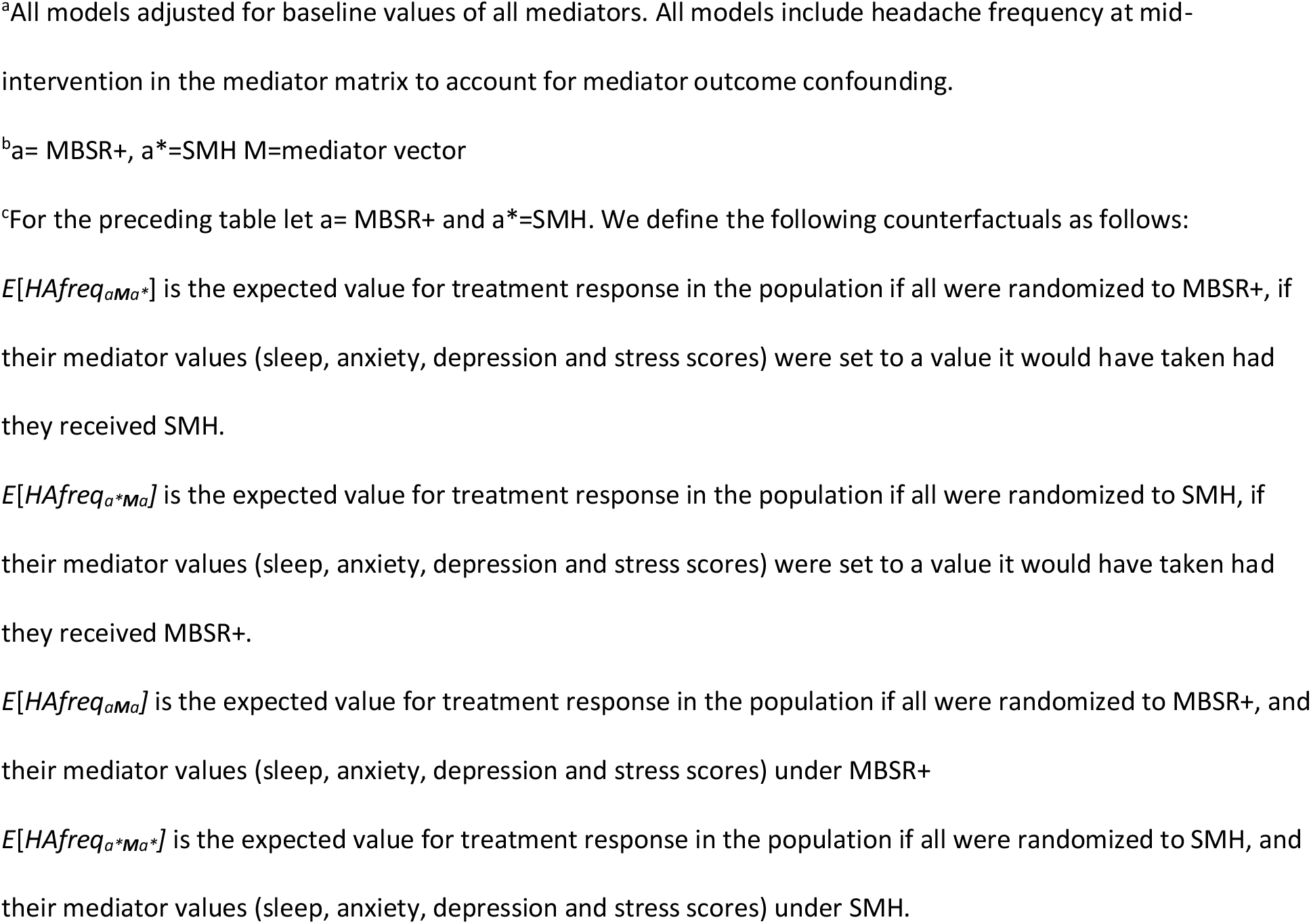
**Counterfactual estimates from the weighting approach to assess mediation effects of anxiety, depression and stress on treatment response**

| Effect<sup>a,b,c</sup> | Estimate | 95% CI |
| --- | --- | --- |
| **Sleep-PSQI** |  |  |
| $E[HAfreq_{aMa*}]$ | 0.42 | **0.38, 0.47** |
| $E[HAfreq_{a*Ma}]$ | 0.28 | **0.24, 0.33** |
| $E[HAfreq_{aMa}]$ | 0.48 | **0.43, 0.54** |
| $E[HAfreq_{a*Ma*}]$ | 0.23 | **0.19, 0.26** |
| **Anxiety-GAD-7** |  |  |
| $E[HAfreq_{aMa*}]$ | 0.45 | 0.41, 0.50 |
| $E[HAfreq_{a*Ma}]$ | 0.28 | 0.25, 0.32 |
| $E[HAfreq_{aMa}]$ | 0.52 | 0.47, 0.56 |
| $E[HAfreq_{a*Ma*}]$ | 0.23 | 0.20, 0.26 |
| **Depression-PHQ-9** |  |  |
| $E[HAfreq_{aMa*}]$ | 0.44 | 0.40, 0.49 |
| $E[HAfreq_{a*Ma}]$ | 0.29 | 0.25, 0.33 |
| $E[HAfreq_{aMa}]$ | 0.52 | 0.47, 0.58 |
| $E[HAfreq_{a*Ma*}]$ | 0.23 | 0.20, 0.26 |
| **Stress-PSS** |  |  |
| $E[HAfreq_{aMa*}]$ | 0.45 | 0.41, 0.50 |
| $E[HAfreq_{a*Ma}]$ | 0.28 | 0.24, 0.32 |
| $E[HAfreq_{aMa}]$ | 0.52 | 0.47, 0.56 |
| $E[HAfreq_{a*Ma*}]$ | 0.23 | 0.20, 0.26 |

<sup>a</sup>All models adjusted for baseline values of all mediators. All models include headache frequency at mid-intervention in the mediator matrix to account for mediator outcome confounding.
<sup>b</sup>a= MBSR+, a\*=SMH M=mediator vector
<sup>c</sup>For the preceding table let a= MBSR+ and a\*=SMH. We define the following counterfactuals as follows:

**Table 6:** Direct and indirect effects of MBSR+ on treatment response.

| Effect<sup>a</sup> | Estimate | 95% CI<sup>b</sup> |
| --- | --- | --- |
| **Sleep** |  |  |
| Direct effect of MBSR | 0.19 | **0.18, 0.20** |
| Indirect effect through sleep | 0.06 | **0.05, 0.08** |
| **Anxiety** |  |  |
| Direct effect of MBSR | 0.22 | **0.21, 0.23** |
| Indirect effect through anxiety | 0.07 | **0.06, 0.08** |
| **Depression** |  |  |
| Direct effect of MBSR | 0.21 | **0.20, 0.22** |
| Indirect effect through depression | 0.08 | **0.07, 0.09** |
| **Stress** |  |  |
| Direct effect of MBSR | 0.22 | **0.21, 0.23** |
| Indirect effect through stress | 0.06 | **0.05, 0.08** |

<sup>a</sup>All models adjusted for baseline values of all mediators. All models include headache frequency at mid-intervention in the mediator matrix to account for mediator outcome confounding.
<sup>b</sup>Confidence intervals are highlighted in bold to identify that direct and indirect effects are significant

## Discussion

We examined the longitudinal relationship between MBSR+ treatment, sleep quality and psychosocial symptoms in the context of a randomized trial using an active control for headache management. We found that sleep quality was significantly improved in both treatment groups at 20 weeks. Though not significant, higher baseline sleep disturbance and psychosocial factors were associated with greater odds of treatment response regardless of treatment condition. The total effect of MBSR+ on treatment response, operationalized as a 50% reduction in headaches at 20 weeks, included indirect effects of the sleep, anxiety, depression and stress (ranging from 6 to 8%) in addition to direct effects of MBSR+.

These results indicate that the effect of MBSR+ extends beyond headache improvement and encapsulates other components of the migraine experience.

The impact of MBSR on psychological well-being is inconsistent and, at best, modest. A meta-analysis of mindfulness studies with specific and non-specific control arms, across several patient groups reported an effect size of mindfulness on anxiety of 0.38 (95% CI 0.12-0.64) and 0.30 (95% CI 0.00-0.59) for depression at 8 weeks, both of which declined by 6 months.^26^ Within pain populations (fibromyalgia and chronic back pain), these effects on depression and anxiety, though favoring mindfulness, were not significant.^26^ Cherkin and colleagues reported significant improvements in depression and anxiety in chronic lower back pain patients randomized to MBSR, cognitive behavioral therapy or usual care up to 26 weeks post intervention.^27^ Effects were seen at 8 weeks post intervention and group differences were most notable comparing MBSR to usual care. A small pilot study assessing MBSR in episodic migraine reported no significant improvements in anxiety, stress, or depression scores.^28^ In our study, group differences between MBSR+ and SMH was −0.13 in anxiety in our migraine patients, which is similar to that observed comparing MBSR and CBT in chronic lower back pain patients at 8 week follow up (-0.18 effect size).^27^ When compared to active treatments, like acceptance based therapy, the effects of MBSR in chronic pain patients on anxiety and depression are small and not significant.^29^ We also find little or no impact of MBSR+ on reports of anxiety or depression in our sample of episodic migraineurs.

We do, however, see effects of treatment on sleep quality, yet no differential benefit of MBSR+ over SMH in reducing sleep disturbance. As with indices of psychological well-being, the impact of MBSR on sleep quality is varied. A systematic review assessing the effects of MBSR on sleep disturbance reported significant improvements in sleep quality in several clinical populations, including fibromyalgia.^30^ Importantly significant improvements were only observed in uncontrolled studies, but was absent in those with an active control arm.^8,30^ Similarly we observed significant improvements in sleep quality but there was no difference between our two active arms at 20 weeks, indicating that both treatments improved sleep. One possibility is that the improvement in sleep quality we observed was related to the reduction in headache frequency, rather than a direct effect of intervention.

We observed significant indirect effects of 6-8%, illustrating that some of the effect of MBSR+ in reducing headache frequency, is mediated through pathways that include reductions in sleep quality, anxiety, depression and stress symptoms. These small mediation effects and the absence of predictive value of baseline scores are likely because our sample generally reported good sleep quality and low anxiety, depression, and stress, leaving little room for improvement. Previous studies have reported that, compared to chronic pain patients with co-morbid psychosocial symptomatology, patients with clinically diagnosed psychiatric disorders report greater symptom reduction after receipt of MBSR.^10-12^ It is plausible that MBSR+ for migraine might be most effective in patients with high baseline levels of these co-morbid factors.

### Strengths and Limitations

This is the largest RCT assessing mindfulness in migraine patients to date, providing the first comprehensive look at MBSR+ in episodic migraine. The comparable active control treatment arm allowed us to draw conclusions that were more precise from analysis. Stratified block randomization mitigated possible imbalances in risk factors between intervention groups and patient retention was excellent. All analyses used ITT, preserving the benefits of randomization. While patients randomized to SMH may have tried alternative stress management techniques or attempted MBSR on their own, the benefits of ITT outweigh the harms and the likelihood of patients trying both treatments is slim given the time burden associated.

However, our study does have some limitations. It is possible the study attracted migraine patients who are not representative of the general migraine population given their interest in non-pharmacological therapy. We recruited from two large health systems and academic campuses, and a large metropolitan community. This limited geographic scope, the demands of weekly and bi-weekly treatment sessions and the requirement to complete imaging likely limit the representativeness of the patient sample.

Through meditation analysis we show that small changes in psychosocial symptoms, though not significant on their own, contributed to treatment response. One drawback of this approach is the inability to disentangle which mediators are responsible for the effect. However, in separate models the effect of each symptom - anxiety, depression, stress, and sleep disturbance - was similar, implying that MBSR+ works through reduction of these negative psychosocial symptoms to reduce headache frequency. There is always the concern of unmeasured confounding especially in light of the strict assumptions which must be employed with these analyses. The randomized design incorporated a rich collection of baseline covariates, diminishing this risk in this study.

## Conclusions

MBSR+ may be a comprehensive therapeutic approach in migraine patients. Benefits may differ based on a patient’s psychological symptom profile and we establish an initial framework which can aid providers in directing patients to appropriate care. Most importantly, MBSR+ provides patients with the opportunity to take responsibility and control over their care eliminating some of the barriers associated with traditional pharmacological options. Patients acquire lifelong skills which once maintained can improve their overall quality of life.

## Data Availability

Data available upon request.

